# The Role of the Emergency Department in Protecting the Hospital as a Critical Infrastructure in the Corona Pandemic Strategies and Experiences of a Rural Sole Acute-Care Clinic

**DOI:** 10.1101/2020.09.07.20185819

**Authors:** S. Kortüm, D. Becker, H.-J. Ott, H-P. Schlaudt

## Abstract

**Background:** The Klinikum Hochrhein is responsible as a regional sole provider for the acute and emergency medical treatment of more than 170.000 people. Against the background of the pandemic spread of SARS-CoV-2 with expected high patient inflows and at the same time endangering one’s own infrastructure due to intraclinical transmissions, the hospital management defined the maintenance of one’s functionality as a priority protection objective in the pandemic. An essential strategic element was a very short-term restructuring of the Emergency Department with the objectives of reducing the number of cases within the clinic, detecting COVID-19 cases as sensitively as possible and separating the patient pathways at an early stage.

**Methods:** The present work is a retrospective analysis of the processes and structures established in the Emergency Department between 27 March 2020 and 20 May 2020. In addition, a retrospective descriptive evaluation of the epidemiological and clinical data of the patients is carried out at the time of first contact during the period mentioned above.

**Results:** After establishing a pre-triage with structured algorithms, all confirmed COVID-19 cases were identified before entering the clinic and assigned to an appropriate treatment pathway. Unprotected entry into hospital structures or nosocomial infections were not observed, although almost 35% of patients with confirmed infection were admitted due to other symptom complexes or injuries. 201 inpatient patients were initially isolated without COVID-19 being confirmed. The number of cases in the Emergency Department was 39% lower than the previous year’s period, thus avoiding crowding.

**Discussion:** The reduction in the number of cases was strategically intended and is primarily the result of a restrictive indication of in-clinical treatment but supported by a decline in emergency consultations that can be noticed anyway. The proportion of false positive triage results is probably dependent on epidemiological activity and was accepted for safety reasons as sufficient resources were available for isolation.

**Conclusion:** Short-term organizational, spatial and procedural restructuring of the Emergency Department has enabled the clinic to achieve its goal of managing the pandemic. The algorithms we developed are particularly well suited to guarantee the desired level of safety in the case of a high pre-test probability.

## Background

The Klinikum Hochrhein is a hospital of advanced care with 9 main departments and supplementary sections with affiliated specialists. The clinic meets the requirements for emergency care in level 2 (extended emergency care). The Emergency Department is run as an independent specialist department and treats approximately 25.000 patient per year with a rate of inpatient admissions of almost 50 %. The nearest hospitals with acute and emergency medical care are not reachable with a travel time of less than 50 minutes, clinics of maximum care not less than 75 minutes.

Due to the geographical and structural conditions, Klinikum Hochrhein is responsible for ensuring emergency care and inpatient treatment for more than 170.000 people as the sole regional provider.

As early as the end of January 2020, the first infections with SARS-CoV-2 were reported in Germany and transmissions by asymptomatic virus carriers were described [1, 2]. Regarding the dynamics of the infection process in Italy, the challenge that was probably faced by hospitals in Germany became clear:

1. Providing clinical resources for a high number of additional acutely ill patients
2. for a prolonged period of time
3. at the same time endangering one’s own functionality through transmissions within the hospital,
4. and at the same time ensuring acute and emergency care for all other (not COVID-19) patients.

A pandemic infection situation means an external and internal hazard over a longer period of time, for which the existing arrangements for a conventional mass casualty incident are insufficient [3]. When developing an appropriate strategy should not be ignored, as outbreaks of SARS-CoV-2 with some fatal consequences have been reported in hospitals and nursing facilities, with asymptomatic virus carriers playing a major role in the infection chains [4-7]. In China, there were approx. 4% of all confirmed cases Healthcare staff [8]. From Italy, a share of about 20% of infected people reported in this group of people [9], which, even after the end of a quarantine period, can still be considered as a source of infection [10].

## Objective and Strategic Focus

**The overarching protection objective was defined by the hospital management as ensuring the acute and emergency medical care of all emergency patients while maintaining the functionality of the hospital**.

In view of the expected high influx of patients, an upstream or parallel infrastructure was established elsewhere as a “buffer zone” for the clarification of the need for treatment, including outpatient smear diagnostics, thus effectively expanding the existing hospital structure [11-15]. However, this strategy puts a strain on the hospital’s limited human, spatial and material resources, which are then unavailable for the original tasks. At the Klinikum Hochrhein, this option had to be discarded at an early stage due to the lack of space reserves in the existing building and the lack of development areas (inner-city location).

Nevertheless, the expected overcrowding of the clinic structures, and in particular of the Emergency Department, entails the risk of mixing infected and non-infected patients and thus of transmissions, including to medical staff [11-13, 16]. Treatments in the core area of the Emergency Department should only be carried out when there is an objective medical need [17].

The hospital management and the crisis unit therefore defined a strategy for the **isolation of the hospital** with the following key elements:

1. Suspension of all elective outpatient and inpatient hospital treatment
2. Limitation of family visits to very few exceptions
3. Close all entrances and use of security personnel, access only with employee ID
4. Use of security personnel on the access for the rescue service
5. Reversing of all patients arriving on foot to an upstream practice with separate access. Only after appropriate indication forwarding to the Emergency Department
6. Reassignment of 2 wards in a separable part of the building with its own access from outside to isolation areas
7. Restriction of outpatient and inpatient treatment to patients who, at the time of presentation, with medical indication need the specific resources of the hospital irrefutable and not deferrable
8. Referring all safely outpatient patients to practicing physicians and/or the health office, including required medical clarification regarding covid-19

The strategic orientation was coordinated with the external partners at district level (district administration, health office, practicing physicians, ambulance service) and largely implemented without problems.

## Restructuring the Emergency Department

The established range of tasks and services of the Emergency Department as the first point of contact and switching point for all acute and emergency patients will remain unrestricted even in the pandemic [18]. However, an additional focus had to be placed on the protection of the functioning of the hospital by preventing unnoticed entry of infections with SARS-CoV-2. After the entrances and elective outpatient units were closed, patients could only enter the hospital via the Emergency Department, which should act as a “second line of medical defense” [19] against the spread of infection. Since an early separation of patient pathways (infectious vs. non-infectious) contributes significantly to the prevention of transmissions [11, 15, 16, 19], the decision on the necessary isolation measures and the necessary level of protection of employees should be made before the patient reaches the building.

Parallel to the establishment of separate inpatient and intensive care areas for confirmed COVID-19 cases, suspected cases and non-COVID-19 patients, 3 **outposts of the Emergency Department** were established:

- The rooms of a practice outside the actual clinic building as a first contact for patients arriving on foot
- A triage place around the ambulance service access for the first contact with lying patients
- Two isolation wards with separate access for the reception and initial care of Covid-19 patients and suspected cases

This “decentralization”, implemented within a few days, meant extensive interventions in the structures and core processes of the Emergency Department, requiring rapid rethinking and high personal commitment by all employees.

### Pre-Triage

The established first assessment (here: Manchester Triage System) had to advance 3 core issues (Pre-Triage) considering the strategic orientation of the clinic:

1. Is the treatment of the patient with the resources of the hospital in the specific situation irrefutable and not deferrable necessary?
2. Is there a suspicion or case of differential diagnostic clarification regarding COVID-19?
3. In which area of the hospital is the first aid for the patient safest, considering infection protection and medical criteria?

### First Experiences

At the beginning of March 2020, a COVID-19 rapid query was first established in the Emergency Department, which was still very closely based on the official case definition published by the RobertKoch-Institute, Berlin, Germany. An identical query could also be agreed with the persons responsible for the ambulance service, so that suspected cases could already be preclinically identified and communicated before arriving at the Emergency Department.

However, the first two cases treated with COVID-19 at the Klinikum Hochrhein were not covered by the official case definition at the time of presentation at the Emergency Department. Transmissions within the clinic could be prevented, but against the background of increasing autochthonous transmissions in the region and the heterogeneous symptoms, neither the clinical nor the epidemiological criteria of the case definition proved to be sufficiently sensitive.

## Case Detection and Patient Allocation

Based on the initial experience, it was decided to develop our own algorithms for case detection and patient allocation. The aim was to achieve the highest possible sensitivity in the detection of COVID-19, while a low specificity with a high number of precautionary isolations was deliberately accepted if inpatient treatment was necessary. Appropriate stationary resources were available due to the overall restrictive indication and could have been extended if necessary.

### Criteria for Case Detection

The **clinical symptoms** of COVID-19 were already described as very heterogeneous in the literature in March 2020. This is all the more true when focusing on the time of first contact in the Emergency Department. The leading symptoms fever, cough and dyspnea are observed in a maximum of 70% of cases [20-23]. In addition, non-specific general symptoms, silent hypoxemia or olfactory disorders are reported even without concomitant respiratory symptoms [23-27]. A significant proportion of patients present themselves asymptomatic at the time of first contact [20].

In addition, patients with COVID-19, whose severity of disease requires hospitalization, often have significant comorbidities that push themselves into the foreground of clinical perception and may lead to misjudgments as to possible infectivity. Also, for admissions on other reasons (e. g. fall or fracture) infection with SARS-CoV-2 as an easily overlooked concomitant disease may occur, which nevertheless requires isolation of the patient.

Screening based solely on **imaging techniques** cannot be recommended [28]. This also applies to **laboratory parameters** available for emergency diagnostics, which are at best indicative and are not immediately available at the time of initial contact [29-31].

A direct **detection of pathogens by means of RT-PCR** is considered to be a diagnostic gold standard [32] but is not of use for pre-triage screening due to the duration of time until the findings are available. The **rapid tests for antibodies** to SARS-CoV-2 available since February 2020 are not suitable for rapid exclusion or for confirming the diagnosis with the necessary safety due to the dynamics of the immune response and the technical test parameters [33, 34].

The **epidemiological history** according to the official case definition (stay in risk areas and/or contact with confirmed cases) can be helpful in the early phase of an outbreak with defined clusters, but with the occurrence of autochthonous transmissions in the respective region, it is no longer sufficiently certain from the aspect of the protective function of the emergency room [13, 35].

**In summary**, a simple one-dimensional screening with high sensitivity is not available at the time of the first contact. In regions with proven autochthonous transmission, it would seem appropriate to handle all patients with symptoms compatible with COVID-19 as suspected cases or as cases of differential diagnostic clarification [36] and to continuously develop its own multimodal algorithms for case detection and patient allocation, taking into account the local framework conditions [12, 19, 37].

### Algorithm for Case Detection

As of March 27, 2020, a specially developed algorithm for case detection was established, which, in addition to broader range of clinical criteria, focuses on the regionally observable epidemiological events in risk facilities, e. g. nursing homes (Figure 1). The expected number of ultimately not confirmed suspected cases was knowingly accepted in order to avoid nosocomial transmissions with the highest possible safety.

**Figure 1:**
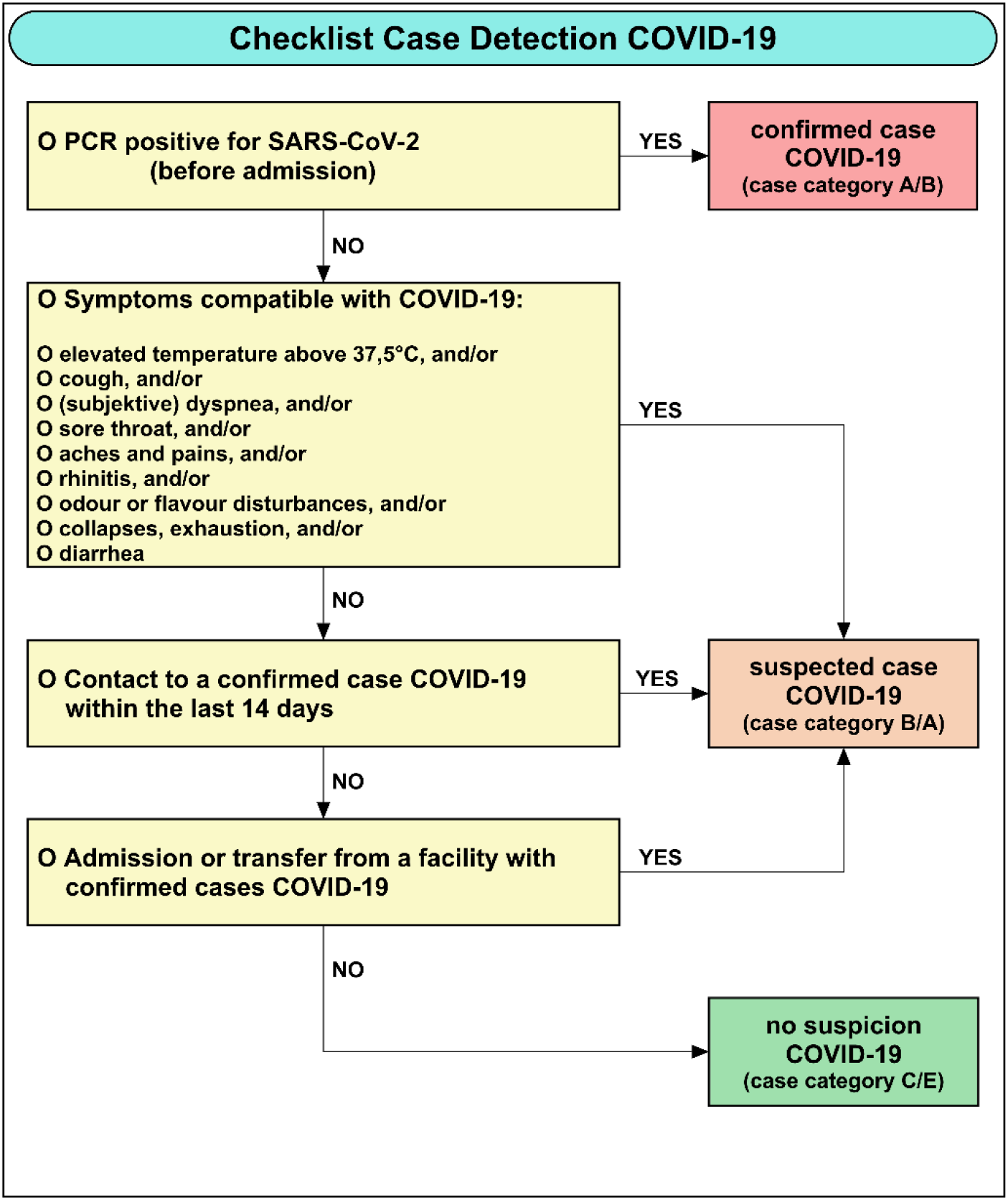
Checklist for the identification of COVID-19 suspected cases in the context of pre-triage

### Case Categories and Patient Pathways

All unplanned incoming patients were assigned 5 defined case categories and allocated spatially separated treatment pathways using a checklist (Figure 2).

**Figure 2:**
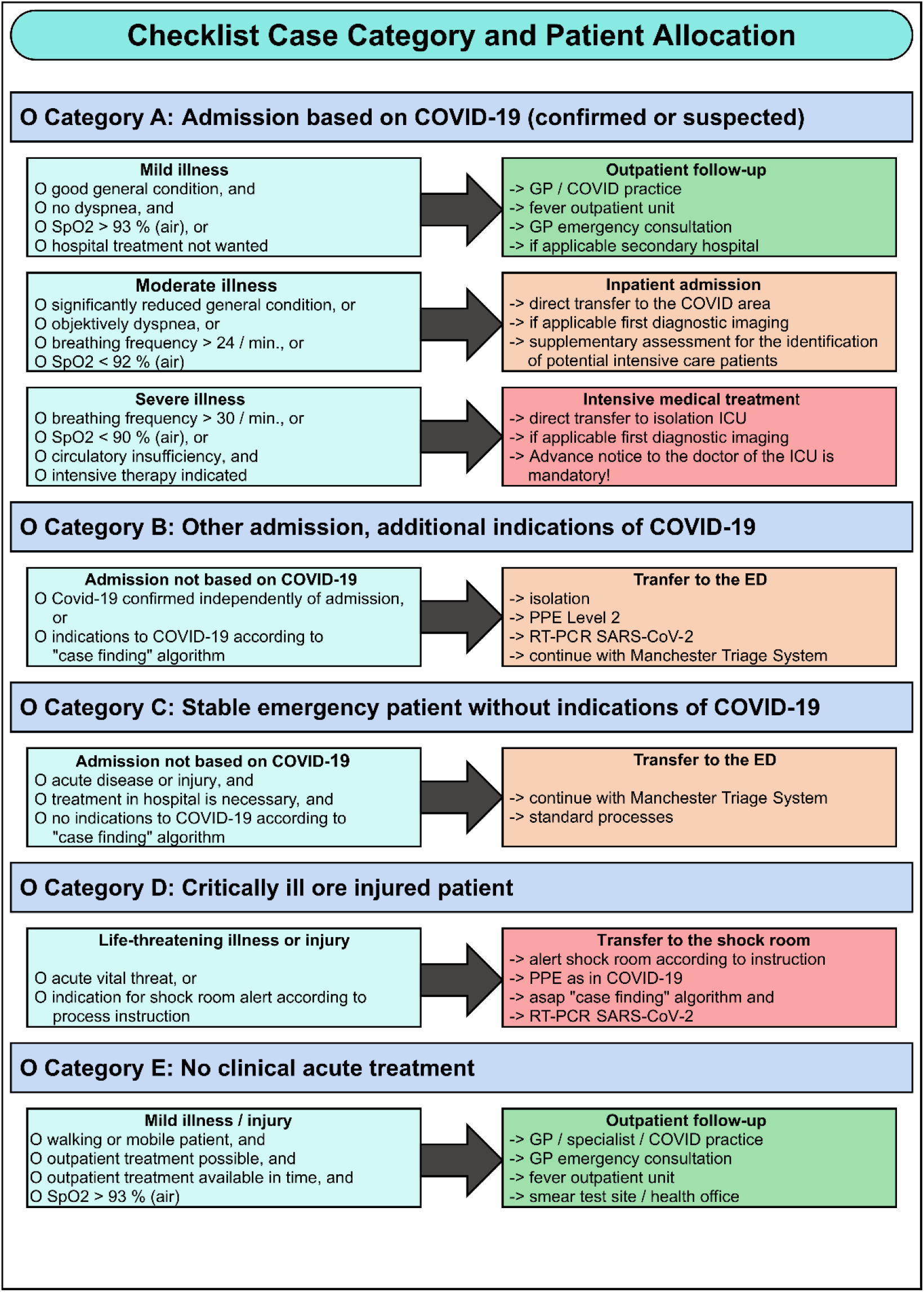
Checklist to define a case category and assignment to a treatment path

Case detection and patient management were carried out by interprofessional triage teams in the context of a pre-triage around the ambulance service access (in the vehicle) or for patients on foot in the practice upstream of the Emergency Department. By clearly defining responsibilities and drawing up special service plans personal protective equipment in high-risk areas could be used very consistently and at the same time in a resource-saving manner. Completely unprotected contacts were avoided by a face mask obligation for all persons within the hospital building.

### Ambulance Service

The ambulance service agreed on a checklist simplified in terms of clinical criteria (Figure 3) in order to identify as many potential COVID-19 patients as possible and to support early and targeted communication at the interface between pre-clinic and Emergency Department.

**Figure 3:**
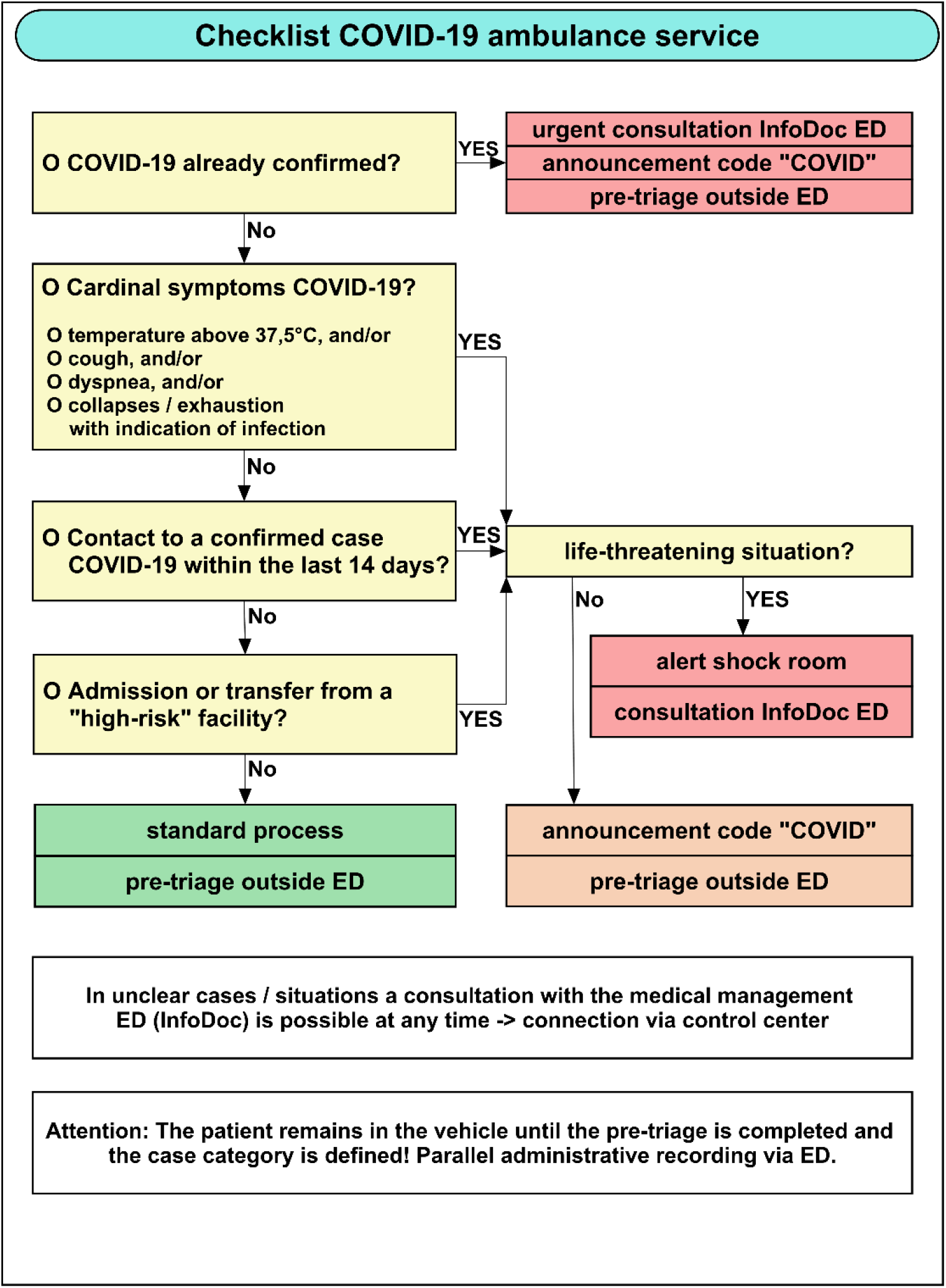
Checklist for early detection of suspected cases in ambulance services

In addition, from 27. March 2020, a doctor from the central emergency room was continuously available to the rescue service as a contact person in cases of doubt (“InfoDoc”), who was alerted via the control center on request. To provide technical support to the InfoDoc, a control center workstation was set up in the Emergency Department at short notice.

## Results

### Case Categories

In the period from 27. March 2020 to 20. May 2020 a total of 2,184 patients were registered in the Emergency Department (previous year period: 3,585). This corresponds to a decrease of almost 39%, which is at least partly due to the consistent focus on irrefutable and not deferrable hospital treatments. All patients were assessed using the case-finding checklist as part of the pre-triage and assigned to a treatment path.

288 of the 2,184 patients (13. 2%) were identified as COVID-19 suspicious and assigned to case category A or B (Figure 4).

**Figure 4:**
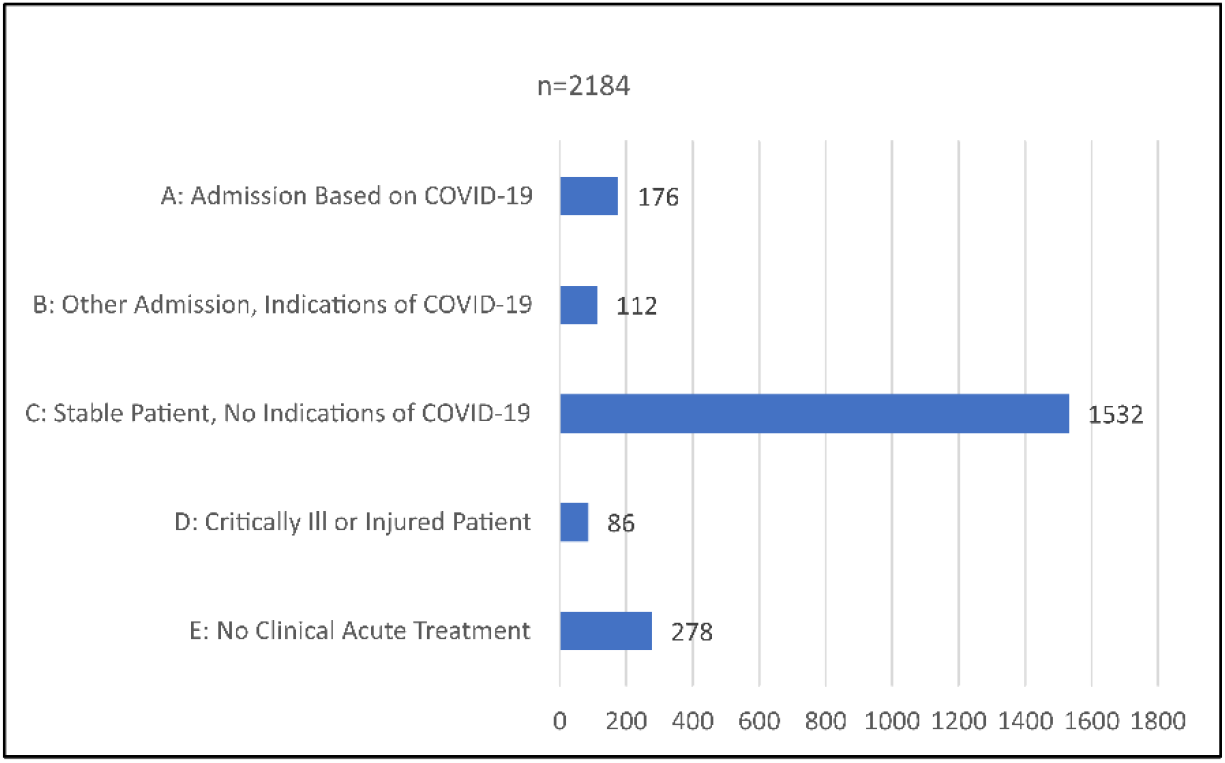
Case Categories of ED patients 27. March to 20. May 2020, n=2184

Of the 288 patients in case categories A and B, 32 (11. 1 %) were discharged into further outpatient treatment after pre-stationary treatment and appropriate information. In 256/288 patients (88. 9%), the clinical indication was given for inpatient admission to one of the isolation wards. The age of inpatient patients ranged from 26 to 97 years, median 78 years.

COVID-19 was confirmed in 55 out of 256 inpatient patients (21. 5%). In case of persistent clinical suspicion despite negative smear test results, isolation was maintained, and the diagnosis was repeated. In 6 patients (10. 9%), the first smear test was negative. In 2 patients (3. 6%), the diagnosis of COVID-19 was finally confirmed only in the third test.

**During the investigation period, all COVID-19 cases confirmed during the inpatient course were correctly identified and isolated as suspicious cases by means of the structured pre-triage. No unnoticed and therefore unprotected entry of the infection into hospital structures and/or nosocomial infections were known**.

### Primary Reasons for Admission

29 of the 55 confirmed Covid-19 disorders (52. 7%) were admitted and announced as a suspected case after the introduction of the “Checklist Ambulance Service”, 7 patients (12. 7%) as “other respiratory problems”. The remaining 19 patients (34. 6%) initially were presented for other reasons in the Emergency Department (Table 1) and would not have been isolated at first without pre-triage.

**Table 1:** Primary Reasons for Admission to the ED in COVID-19 Patients Confirmed in the Course, n=55

|  |  |  |
| --- | --- | --- |
| Anlass | n | % |
| COVID-19 (Suspected or Confirmed) | 29 | 52,7 |
| Other Respiratory Problems | 7 | 12,7 |
| Gastrointestinal Problems | 4 | 7,3 |
| Collapse / Syncope | 4 | 7,3 |
| Reduced General Condition / Exsiccosis | 3 | 5,5 |
| Cardiac Disease | 2 | 5,5 |
| Neurological Disorder | 2 | 3,6 |
| Injury after Fall | 2 | 3,6 |
| Urinary Infection | 1 | 1,8 |
| Hyperglycaemia | 1 | 1,8 |

### Symptoms at First Contact

56. 4% (31/55) of the confirmed COVID-19 cases had elevated temperature above 37. 5°C at the time of first contact, 61. 8% (34/55) cough and 60. 0% (33/55) dyspnea (Figure 5).

**Figure 5:**
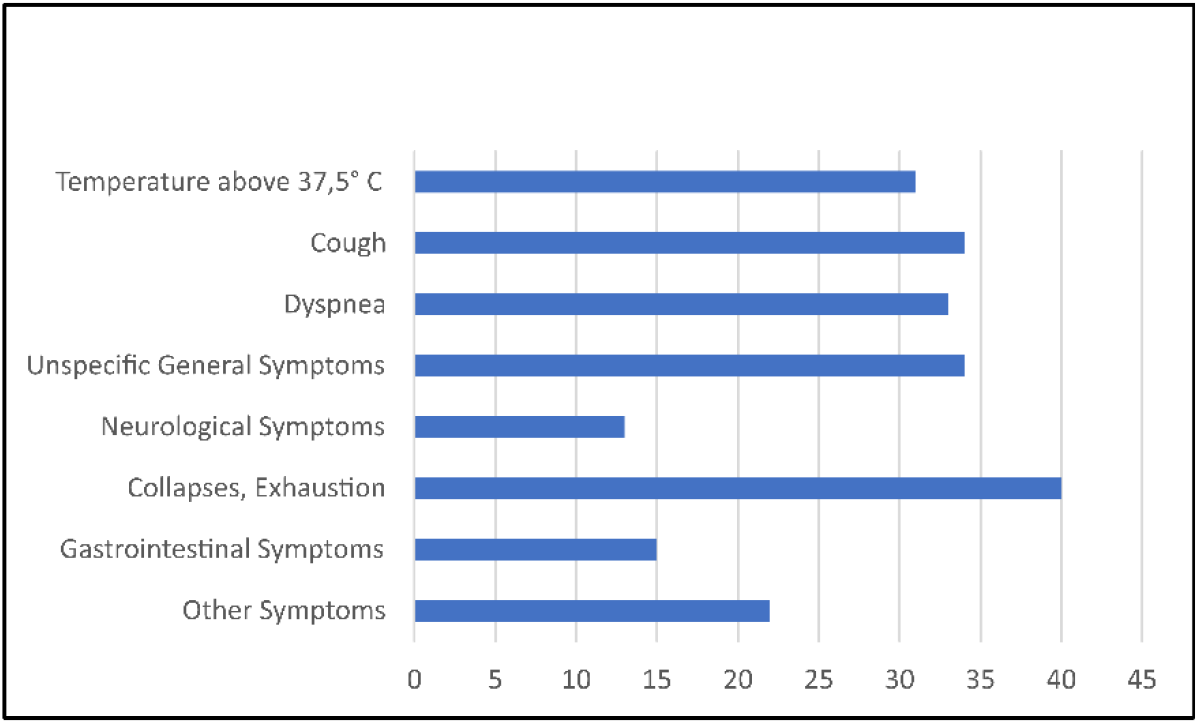
Clinical Symptoms of Confirmed COVID-19 Cases at the Time of First Contact, n=55

61. 8% (34/55) of the patients had non-specific general symptoms (throat pain, limb pain, cold), 23. 6% (13/55) had neurological symptoms (dizziness, reduced vigilance, anosmia). In 72. 7% (40/55) of the cases, collapses and/or exhaustion were reported, in 27. 3% (15/55) gastrointestinal problems (diarrhea, vomiting).

Considering the intended reliable and early identification of cases, it is significant that 21. 8% (12/55) of patients did not show any respiratory symptoms at the time of first contact. 12. 7% (7/55) had neither respiratory symptoms nor fever. Unspecific worsening of the general condition and the tendency to collapse were the focus of the clinical picture in these – often very old – patients.

### Epidemiological History

In 29 (52. 7%) of the patients, neither direct contact with confirmed COVID-19 cases nor a stay in high-risk areas or facilities could be recorded at the time of pre-triage (Figure 6).

**Figure 6:**
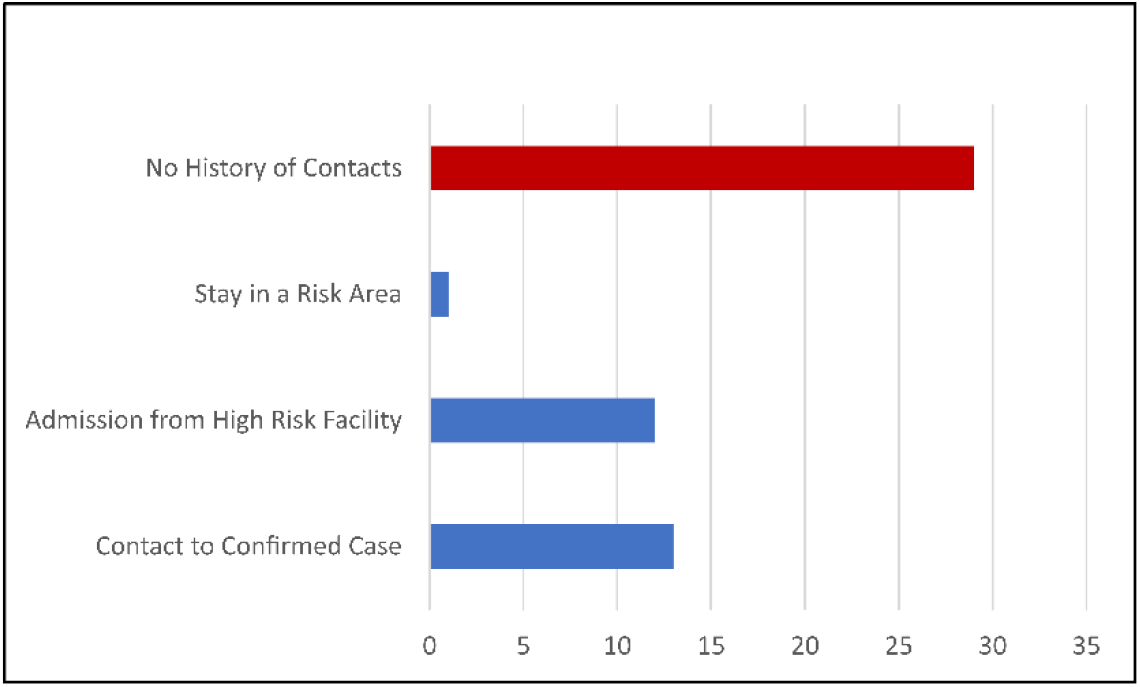
Epidemiological History of Confirmed COVID-19 Cases, n=55

Only 1 patient had been in one of the officially listed risk areas prior to the disease. 4 out of 7 (57. 1 %) of patients with only non-specific general symptoms were correctly identified as suspected case due to the admission from a risk facility.

## Discussion

The primary objective of maintaining acute clinical care for all patients (COVID-19 and non-COVID-19) was achieved according to the strategic orientation of the clinic. At this point, the Emergency Department has the key function of identifying those patients who need treatment with the hospital resources for medical reasons and referring other cases to the outpatient sector. At the same time, COVID-19 (suspected) cases must be identified with the highest possible sensitivity in order to prevent an unnoticed spread within the hospital safely.

In order to fulfil this task, extensive organizational, spatial and process adjustments had to be implemented in the shortest possible time in the daily operation of the Emergency Department, which required a high degree of flexibility from all parties involved.

The reduction in the number of cases observed during the investigation period of almost 39% compared to the previous year’s period was strategically intended to prevent the overloading of clinical structures feared according to the early reports from Italy and France. It is primarily the result of a restrictive indication of in-clinical treatment, but undoubtedly favored by a decline in emergency consultations also observed elsewhere.

This procedure also explains the very low proportion of suspected cases with only outpatient or pre-inpatient treatment. Pure “work-up examinations” were not carried out by us in consultation with the competent health authority at any time. The overall concept was only successful because a close cooperation and coordination with the outpatient structures (corona consultation hours, fever outpatient unit, smear test site, telephone hotline) was reliably possible.

The algorithms for case detecting and patient allocation have proven to be effective under our framework conditions. All confirmed COVID-19 cases were correctly identified in the structured pre-triage, unprotected contacts with employees or other patients did not take place, nosocomial transmissions were not known. 20 % of our patients had neither respiratory symptoms nor contact with COVID-19 cases, not even in the context of a recognizable outbreak, and would not have been immediately recognized if the “official” case criteria had been applied.

Almost 35% of patients with COVID-19 in our patient population were admitted primarily due to other symptoms or injuries. These may obscure and/or distract evidence of an infectious event, which underlines the need for a highly sensitive case detection.

The high number of false positive results from the pre-triage is not satisfactory, but was accepted in favor of reliable case detection to avoid unnoticed transmissions within the hospital. This presupposes that the appropriate clinical resources for the isolation of patients are actually available within the framework of an overall concept.

In this context, it should also be noted that the proportion of incorrectly positive triage results is not constant, but appears to depend on the epidemiological activity (pre-test probability) and was significantly lower at the beginning of the investigation period. Further analyses and a prospective re-evaluation of the algorithms are planned.

The very heterogeneous epidemiological data of our patients reflect an already pronounced – partly subclinical – autochthonous transmission of the infection. Stays in risk areas played a completely subordinate role in our patient population, not least because of demographic data (very old people). The definition of risk facilities when admitting or relocating patients, considering the regional course of infection, has proven to be effective.

As a limiting factor, it should be noted that our strategic approach has been successful in the context of the regional framework conditions and the structure of the hospital and cannot therefore be transferred uncritically to other areas. Due to the influence of the pre-test probability, the developed algorithms are particularly suitable for use in times of high epidemiological activity with autochthonous transmissions. Due to the relatively low number of cases so far, further evaluation is necessary and planned.

## Conclusion

Through short-term organizational, spatial and procedural restructuring of the Emergency Department, the clinic’s goal of coping with the pandemic was achieved. The algorithms we have developed are particularly well suited to ensure the desired level of security when there is a high probability of pre-testing.

## Data Availability

All mentioned data are available from the authors

## Compliance With Ethical Guidelines

**Conflict of interest**. S. Kortüm, D. Becker, H. -J. Ott and H. -P. Schlaudt state that there is no conflict of interest.

**No studies on humans or animals** were conducted by the authors for this paper. The quoted studies are subject to the respective ethical guidelines.

